# Temporal association of reduced serum vitamin D with COVID-19 infection: A single-institution case-control and historical cohort study

**DOI:** 10.1101/2021.06.03.21258330

**Authors:** Diviya Gupta, Sahit Menon, Michael H. Criqui, Bryan K. Sun

## Abstract

**Objectives:** Vitamin D supplementation has been proposed for the prevention and treatment of COVID-19, but the relationship between the two is inconclusive: Reduced serum vitamin D may predispose to COVID-19, but it may also be a secondary consequence of infection. The objective of this study was to assess the temporal association between serum vitamin D levels and COVID-19.

**Design:** A single-institution case-control study and a historical cohort study

**Setting:** University of California San Diego (UCSD) Health System in San Diego, California

**Participants:** Patients testing positive for COVID-19 from January 1, 2020 to September 30, 2020 with serum 25-hydroxy-vitamin D (25(OH)D) measured within 180 days of diagnosis. Patients were separated based on whether 25(OH)D was measured before (n=107; “pre-diagnosis”) or after (n=203; “post-diagnosis”) COVID-19 diagnosis. Subjects with 25(OH)D assessments prior to COVID-19 diagnosis were evaluated using a case-control study design, while subjects with 25(OH)D measured after COVID-19 diagnosis were analyzed with a historical cohort study design.

**Primary and Secondary Outcome Measures:** In the pre-diagnosis study, a conditional logistic regression was performed using COVID-19 infection status as the binary dependent variable. In the post-diagnosis study, an ordinary least squares regression was performed using serum 25(OH)D levels as the continuous dependent variable.

**Results:** Serum 25(OH)D levels were not associated with the odds of subsequently testing positive for COVID-19 (OR 1.00, 95% CI: 0.98 to 1.02, p = 0.982). However, COVID-19 positive individuals had serum 25(OH)D measurements that were lower by 2.70 ng/mL (95% CI: −5.19 to −0.20, p = 0.034) compared to controls.

**Conclusions:** In our study population, serum 25(OH)D levels were not associated with risk of testing positive for COVID-19 but were reduced in subjects after being diagnosed with COVID-19 infection. These results raise the possibility that reduced serum 25(OH)D may be a consequence and not a cause of COVID-19 infection.

## INTRODUCTION

SARS-CoV-2, the coronavirus that causes COVID-19, has claimed over 3.5 million lives globally. Although there is optimism that vaccination efforts will eventually bring the pandemic under control, there has been widespread interest in complementary measures to mitigate the risk of viral infection and mortality. Vitamin D, available as an inexpensive supplement, gained extensive attention for its potential role in the prevention and treatment of COVID-19. However, the relationship between vitamin D and COVID-19 is unclear.

Mechanistically, vitamin D may enhance the immune response to SARS-CoV-2 in several ways. Vitamin D boosts innate immunity by augmenting the expression of human cathelicidin antimicrobial peptide (CAMP)^1^ in lung and skin epithelial cells^2^, which has been shown to attenuate the infectivity and viability of viruses^3^. In addition, vitamin D prevents excessive adaptive immune responses^4^. Because systemic inflammatory responses have been associated with respiratory distress and mortality in patients with severe COVID-19^5^, it has been proposed that vitamin D supplementation could mitigate these harmful inflammatory reactions.

Some medical experts and public figures have recommended vitamin D supplementation for COVID-19^6^, using serum 25-hydroxy-vitamin D (25(OH)D), a standard laboratory serum marker of vitamin D stores, as a primary biomarker for vitamin sufficiency. Recommendations for supplementation have particularly been endorsed for populations with the highest COVID-19 risk who also have higher rates of vitamin D deficiency, including the elderly, individuals with chronic diseases, darker-complected individuals such as African Americans, and those who live at high latitudes^7^.

The studies that have evaluated the correlation between serum vitamin D and COVID-19 have shown mixed results. Some reports have concluded that there is an association between vitamin D deficiency and increased susceptibility to COVID-19 infection^8,9^, but others have not^10,11^. Other studies, focusing on different endpoints, have found that 25(OH)D deficiency is correlated with a higher risk of intensive care unit admission, ventilation dependency, and survival rate^12–16^. Beneficial outcomes from vitamin D supplementation trials have been reported^17^, although these therapeutic studies have not always been done in randomized groups^18^, and have had modest sample sizes. It has been postulated that any beneficial effect of vitamin D on severe COVID-19 could be masked by the effect of other adjunctive treatments such as dexamethasone^19^. Highly powered, randomized controlled trials will be needed to definitively test for causality.

The differences in conclusions among published results may be caused, at least in part, by significant methodological differences. Some reports have examined serum 25(OH)D on the basis of country-wide averages^20,21^ or have inferred vitamin D status based on geographic latitude^22^, and did not directly assess serum vitamin D in individual subjects. Other studies have used vitamin D results that were measured years prior to COVID-19 testing^23^, which may have resulted in inaccurate representations because 25(OH)D levels can change significantly with time^24^ and season^25^. Finally, some reports assessed 25(OH)D levels drawn prior to COVID-19 diagnosis^8,9^, others relied on measurements taken after diagnosis^12,13^, and others included both^26^. Because severe illness itself can cause rapid reduction of serum 25(OH)D^27^, the timing of laboratory measurement is critical: Vitamin D deficiency may predispose to COVID-19, but it is also possible that COVID-19 infection can reduce 25(OH)D as well.

In this report, we sought to address these methodological differences in examining the temporal correlation between serum 25(OH)D and COVID-19 infection. We performed two complementary single-center studies examining patients testing positive for COVID-19 between January 1, 2020 to September 30, 2020 in the University of California San Diego (UCSD) Health system who had a serum 25(OH)D assessment within 180 days of diagnosis. These dates capture the vast majority of COVID-19 cases in the UCSD Health system from the onset of the COVID-19 pandemic until the initiation of vaccinations.

We evaluated the study populations with complementary approaches: In the first study, we used a case-control design to compare 25(OH)D levels drawn prior to COVID-19 positive diagnosis against COVID-19 negative controls matched by age, sex, body mass index (BMI), diabetes, hypertension, time from vitamin D draw, and season that the 25(OH)D test was performed. In the second study, we applied a cohort study design to assess serum 25(OH)D levels drawn after COVID-19 diagnosis, applying the same matching criteria. Finally, we performed a subgroup analysis of the second study to specifically examine COVID-19 patients whose disease was severe enough to require hospitalization, comparing them against a matched hospitalized cohort that was COVID-19 negative.

Our overarching approach was based on the reasoning that if vitamin D deficiency increased susceptibility to COVID-19 infection, then serum 25(OH)D levels drawn prior to diagnosis would be significantly lower in cases vs. controls. However, if 25(OH)D was lower in COVID-19 cases only if measured after diagnosis, then COVID-19 infection itself may have led to reduction in vitamin D levels. The null hypotheses were that there was no correlation between 25(OH)D levels and COVID-19 in either study. Finally, by focusing on the subgroup of patients requiring hospitalization for COVID-19 and comparing it to a matched hospitalized cohort, we sought to examine if any correlation between 25(OH)D and COVID-19 was amplified among patients affected by severe forms of COVID-19 illness.

## RESULTS

### Serum 25(OH)D was lower in COVID-19 subjects tested after but not before diagnosis

Baseline characteristics of the study cohorts are shown in **Table 1**. Covariates consisted of established risk factors for COVID-19 susceptibility^28^ that included age, sex, obesity, and medical comorbidities. In addition, due to seasonal variation in serum vitamin D levels, control subjects were matched by meteorological season that serum vitamin D was measured, as well as length of delay between serum vitamin D assessment and COVID-19 test. Matched study populations were balanced between groups.

**Table 1.** Characteristics of COVID-19 study population and controls. Continuous covariates are reported as mean ± standard deviation (SD) and compared using unpaired t-tests. Categorical variables are reported as numbers (no.) and percentages (%) and compared using Chi-square tests. A p-value < 0.05 was considered a significant difference for covariates. Season dates were defined by meteorological seasons within the study interval.

| Characteristic | Serum 25(OH)D measured before COVID-19 diagnosis |  |  | Serum 25(OH)D measured after COVID-19 diagnosis |  |  |
| --- | --- | --- | --- | --- | --- | --- |
|  | *COVID-19 positive*<br>*(n=107)* | *COVID-19 negative*<br>*(n=214)* | *p-value* | *COVID-19 positive*<br>*(n=203)* | *COVID-19 negative*<br>*(n=406)* | *p-value* |
| Age (years) ± SD | 51.24 ± 15.99 | 52.55 ± 15.31 | 0.486 | 52.66 ± 15.73 | 53.35 ± 15.19 | 0.604 |
| Sex – no. (%) |  |  |  |  |  |  |
| Male | 53 (49.53) | 106 (49.53) | 1 | 123 (60.59) | 246 (60.59) | 1 |
| Female | 54 (50.47) | 108 (50.47) |  | 80 (39.41) | 160 (39.41) |  |
| Body Mass Index ± SD | 27.80 ± 5.77 | 27.51 ± 5.43 | 0.665 | 28.03 ± 6.49 | 27.61 ± 5.93 | 0.438 |
| Diabetes – no. (%) |  |  |  |  |  |  |
| Yes | 22 (20.56) | 44 (20.56) | 1 | 70 (34.48) | 140 (34.48) | 1 |
| No | 85 (79.44) | 170 (79.44) |  | 133 (65.52) | 266 (65.52) |  |
| Hypertension – no. (%) |  |  |  |  |  |  |
| Yes | 50 (46.73) | 100 (46.73) | 1 | 109 (53.69) | 218 (53.69) | 1 |
| No | 57 (53.27) | 114 (53.27) |  | 94 (46.31) | 188 (46.31) |  |
| Days between vitamin D and COVID-19 tests ± SD | 77.39 ± 39.88 | 77.67 ± 39.61 | 0.953 | 32.28 ± 40.40 | 32.33 ± 38.89 | 0.989 |
| Season – no. (%) |  |  |  |  |  |  |
| Winter (January 1 – February 29) | 36 (33.64) | 72 (33.64) | 0.981 | - | - | 0.984 |
| Spring (March 1 – May 31) | 33 (30.84) | 68 (31.78) |  | 42 (20.69) | 82 (20.20) |  |
| Summer (June 1 – August 31) | 38 (35.51) | 74 (34.58) |  | 124 (61.08) | 251 (61.82) |  |
| Fall (September 1 – September 30) | - | - |  | 37 (18.23) | 73 (17.98) |  |

For subjects in which 25(OH)D serum levels were assessed prior to COVID-19 test using a case-control design, the mean serum 25(OH)D was 35.45 ng/mL (SD 13.74) for cases and 35.41 ng/mL (SD 13.81) for controls. A one-unit increase in serum 25(OH)D did not affect the odds of contracting COVID-19 (OR 1.00, 95% CI 0.98 to 1.02, p = 0.98). These data revealed no significant association between serum 25(OH)D and the odds of subsequent COVID-19 positivity.

By contrast, in subjects for whom 25(OH)D was measured after diagnosis using the cohort design, subsequent assessment of 25(OH)D showed a mean serum 25(OH)D of 30.46 ng/mL (SD 15.50) for cases and 33.16 ng/mL (SD 15.72) for controls. COVID-19 positivity was associated with serum 25(OH)D levels that were lower by 2.70 ng/mL on average (95% CI −5.19 to −0.20, p = 0.03) (**Table 2**). These data indicated that COVID-19 positive subjects showed a significant reduction in 25(OH)D compared to matched COVID-19 negative subjects.

**Table 2.** Associations of serum 25-hydroxy vitamin D with COVID-19 infection. Serum 25(OH)D for cases and controls are reported as mean levels in ng/mL ± standard deviation (SD). In the pre-diagnosis study, a conditional logistic regression was performed in which the independent variable is continuous (i.e., serum 25(OH)D level) and the dependent variable is binary (i.e., COVID-19 infection status). In the post-diagnosis study, an ordinary least squares regression was performed, in which the independent variable is binary (i.e., COVID-19 infection status) and the dependent variable is continuous (i.e., serum 25(OH)D level).

| **Pre-Diagnosis –**<br>**Conditional Logistic Regression** |  |  |  |  |  |
| --- | --- | --- | --- | --- | --- |
| **Predictor** | **Cases (N=107)**<br>**Serum 25(OH)D**<br>**(ng/mL) ± SD** | **Controls (N=214)**<br>**Serum 25(OH)D**<br>**(ng/mL) ± SD** | **Odds Ratio** | **95% CI** | **p-value** |
| Vitamin D (ng/mL) | 35.45 ± 13.74 | 35.41 ± 13.81 | 1.00 | 0.98 to 1.02 | 0.982 |
| **Post-Diagnosis –**<br>**Ordinary Least Squares Regression** |  |  |  |  |  |
| **Predictor** | **Cases (N=203)**<br>**Serum 25(OH)D**<br>**(ng/mL) ± SD** | **Controls (N=406)**<br>**Serum 25(OH)D**<br>**(ng/mL) ± SD** | **Beta**<br>**Estimate** | **95% CI** | **p-value** |
| COVID-19 Infection | 30.46 ± 15.50 | 33.16 ± 15.72 | -2.696 | -5.19 to -0.20 | 0.034* |

Abbreviations: CI = confidence interval; 25(OH)D = 25-hydroxy vitamin D
\*significant at $p < 0.05$

### Reduced 25(OH)D in COVID-19 positive hospitalized patients compared to a COVID-19 negative hospitalized cohort

To assess for potential correlation between serum 25(OH)D and severe COVID-19 infection, a subgroup analysis was performed to compare COVID-19 subjects requiring hospitalization against a COVID-negative hospitalized control group. To promote matching of patients with comparable disease severity, control patients were additionally matched by length of hospital stay, an indicator of disease severity^29^. Baseline characteristics of the matched analysis for the hospitalized cohort are shown in **Table 3**. No significant differences were observed for matching characteristics between cases and controls.

**Table 3.** Characteristics of hospitalized COVID-19 study population and controls. Continuous covariates are reported as mean ± standard deviation (SD) and compared using unpaired t-tests. Categorical variables are reported as numbers (no.) and percentages (%) and compared using chi-square tests. A p-value less than 0.05 was considered a significant difference for covariates. Season dates were defined by meteorological seasons within the study interval.

| Characteristic | COVID-19 positive<br>(n=120) | COVID-19 negative<br>(n=240) | p-value |
| --- | --- | --- | --- |
| Age (years) ± SD | 55.86 ± 14.60 | 57.85 ± 14.03 | 0.218 |
| Sex – no. (%) |  |  |  |
| *Male* | 82 (68.33) | 165 (68.75) | 0.936 |
| *Female* | 38 (31.67) | 75 (31.25) |  |
| Body Mass Index ± SD | 28.12 ± 6.79 | 26.77 ± 6.28 | 0.069 |
| Diabetes – no. (%) |  |  |  |
| *Yes* | 59 (49.17) | 115 (47.92) | 0.823 |
| *No* | 61 (50.83) | 125 (52.08) |  |
| Hypertension – no. (%) |  |  |  |
| *Yes* | 83 (69.17) | 165 (68.75) | 0.936 |
| *No* | 37 (30.83) | 75 (31.35) |  |
| Days between vitamin D test and<br>COVID-19 hospitalization<br>± SD | 21.02 ± 34.39 | 19.90 ± 29.99 | 0.763 |
| Season – no. (%) |  |  |  |
| *Winter (January 1 – February 29)* | 7 (5.83) | 16 (6.67) | 0.972 |
| *Spring (March 1 – May 31)* | 37 (30.83) | 77 (32.08) |  |
| *Summer (June 1 – August 31)* | 67 (55.83) | 128 (53.33) |  |
| *Fall (September 1 – September 30)* | 9 (7.50) | 19 (7.92) |  |
| Length of Hospitalization (days) ±<br>SD | 21.37 ± 21.43 | 17.20 ± 17.80 | 0.071 |

Within a 180-day window following inpatient admission, COVID-19 positive hospitalized cases had a mean 25(OH)D of 23.92 ng/mL (SD 13.45), while COVID-19 negative hospitalized controls had a mean 25(OH)D level of 27.26 ng/mL (SD 15.35). Patient hospitalization due to COVID-19 was associated with 3.34 ng/mL (95% CI −6.30 to −0.38, p = 0.027) lower serum 25(OH)D levels compared to hospitalized COVID-19 negative patients (**Table 4**). These data indicated that COVID-19 positive hospitalized subjects showed a significant reduction in 25(OH)D compared to COVID-19 negative hospitalized subjects.

**Table 4.** Association of serum 25-hydroxy vitamin D with severe COVID-19 infection. Serum 25(OH)D for cases and controls are reported as mean levels in ng/mL ± standard deviation (SD). An ordinary least squares regression was performed in which the independent variable is binary (i.e., hospitalization due to COVID-19 or hospitalization due to another cause) and the dependent variable is continuous (i.e., serum 25(OH)D level).

| Ordinary Least Squares Regression |  |  |  |  |  |
| --- | --- | --- | --- | --- | --- |
| Predictor | Cases (N=120)<br>Serum 25(OH)D<br>(ng/mL) ± SD | Controls (N=240)<br>Serum 25(OH)D<br>(ng/mL) ± SD | Beta<br>Estimate | 95% CI | p-value |
| COVID-19 Hospitalization | 23.92 ± 13.45 | 27.26 ± 15.35 | -3.340 | -6.30 to -0.38 | 0.027* |

Abbreviations: CI = confidence interval; 25(OH)D = 25-hydroxy vitamin D
\*significant at $p < 0.05$

## DISCUSSION

Defining the temporal association between serum vitamin D and COVID-19 infection provides an important basis for evaluating the potential use of vitamin D supplementation to prevent COVID-19 infection and/or mitigate disease severity. Our single-center study found that COVID-19 infection and hospitalization were associated with lower serum vitamin D levels drawn after diagnosis or hospital admission. However, serum 25(OH)D levels did not affect the odds of initially testing positive for COVID-19, indicating that lower vitamin D was not a risk factor for COVID-19 infection in our study population. Among published studies, some reports have proposed that vitamin D insufficiency, defined as serum concentrations 20-30 ng/mL, or vitamin D deficiency, defined as concentrations <20 ng/mL, may be risk factors for COVID-19 infection^30^. To examine this possibility, we also stratified our pre-diagnosis subjects into these categories. We found no statistically significant association with increased odds of COVID-19 with either vitamin D insufficiency or deficiency compared to the sufficient (>30 ng/mL) reference group (**Supplemental Table 1)**. Viewed together, our results are consistent with other reports that identified lower 25(OH)D in association with COVID-19^8,9,12–14,16^, but raise the possibility that lower 25(OH)D levels may be an outcome of COVID-19 infection rather than a cause of it.

These findings are consistent with prior observations that have examined the relationship between vitamin D and other respiratory illnesses. While vitamin D deficiency has been associated with increased risk of respiratory infections^31^, acute illness itself can also reduce serum 25(OH)D through fluid shifts, depletion of serum binding proteins, and renal wasting^27^. Acute inflammation following surgery has been associated with a reduction of serum 25(OH)D within 48 hours^32^. Inflammatory mediators increase the activity of CYP24A1 and CYP27B1, enzymes that metabolize vitamin D pathway compounds^33^. Vitamin D metabolism is dysregulated in patients with asthma and chronic obstructive pulmonary diseases, leading some investigators to suggest that the relationship between airway inflammation and vitamin D deficiency is bidirectional^34^. These findings support the biological plausibility that elevated rates of vitamin D deficiency observed in subjects infected by COVID-19 could be, at least in part, secondary to the infection itself^35^.

Our study has several strengths. First, our data from a single institution directly assessed 25(OH)D and COVID-19 laboratory results using uniform, standardized assays, minimizing the significant variations that can occur between different testing methodologies^36^. Second, our restriction of 25(OH)D values to a 180-day window, matching to controls by season, and matching by time interval between 25(OH)D and COVID-19 test addresses important time-dependent effects that affect 25(OH)D levels^23,25^. Third, our stratification of 25(OH)D data from the same institutional population into subjects who were tested before vs. after COVID-19 diagnosis allowed us to specifically assess the temporal relationship between vitamin D and COVID-19.

Our study also has several limitations. The retrospective design does not allow for determination of causality, and our modest sample size is not powered to detect smaller but potentially significant correlations. Additionally, although we matched baseline characteristics between cases and controls, other unaccounted confounders could have affected the results. In particular, data on the racial identity of our study subjects was incomplete, which did not allow us to include this factor in cohort matching. Racial disparities in COVID-19 illness have been clearly established, with one retrospective study finding that a positive COVID-19 test was significantly associated with lower vitamin D levels in Black individuals, but not with lower vitamin D levels in White individuals^38^. Our health system’s general catchment area (San Diego county) has a lower Black population (∼5.5%) than the U.S. national average (∼13.4%). Therefore, the results from our analysis may differ from other study populations, and do not argue against the potential utility of Vitamin D supplementation for specific populations.

Ultimately, intervention trials will provide the most conclusive insight to the therapeutic value of vitamin D supplementation for COVID-19. Early reports have shown mixed results: A small (n=76) randomized trial indicated that oral calcifediol supplementation reduced the need for intensive care unit admission in COVID-19 infected subjects^17^, though the trial was not placebo-controlled and did not measure baseline or post-treatment serum vitamin D levels. By contrast, a randomized, double-blind placebo-controlled trial on hospitalized COVID-19 patients found no benefit to a single 200,000 IU dose of vitamin D3 on the length of hospital stay^39^. Viewed together with the results from our study and taken in context with other published studies to date, we recommend caution in the therapeutic expectations for Vitamin D supplementation in the prevention and/or treatment of COVID-19.

## METHODS

### Study Population

The UCSD Institutional Review Board approved this study and approved a waiver for informed consent based on the requirements outlined in the Code of Federal Regulations on the Protection of Human Subjects (45 CFR 46). All data collection and analysis were performed in accordance with relevant guidelines and regulations. Data was collected for all subjects tested for COVID-19 through the UCSD Healthcare system from January 1, 2020 to September 30, 2020 (n=6,050), capturing an interval from the onset of local infections to the time prior to initiation of COVID-19 vaccinations.

For primary analysis **(Figure 1)**, cases were identified as patients who had serum 25(OH)D drawn within 180 days of COVID-19 diagnosis (n=346). Seventeen patients had serum 25(OH)D drawn within 14 days of COVID-19 diagnosis and were excluded to account for the SARS-CoV-2 incubation period and minimize confounding by possible early manifestations of COVID-19^40^. Additionally, 18 patients in the pre-diagnosis study received vitamin D supplementation after serum 25(OH)D testing but before COVID-19 diagnosis; these subjects were excluded due to probable changes in serum 25(OH)D levels from supplementation. One patient was removed from the pre-diagnosis study due to incomplete data on matching covariates.

**Figure 1.**
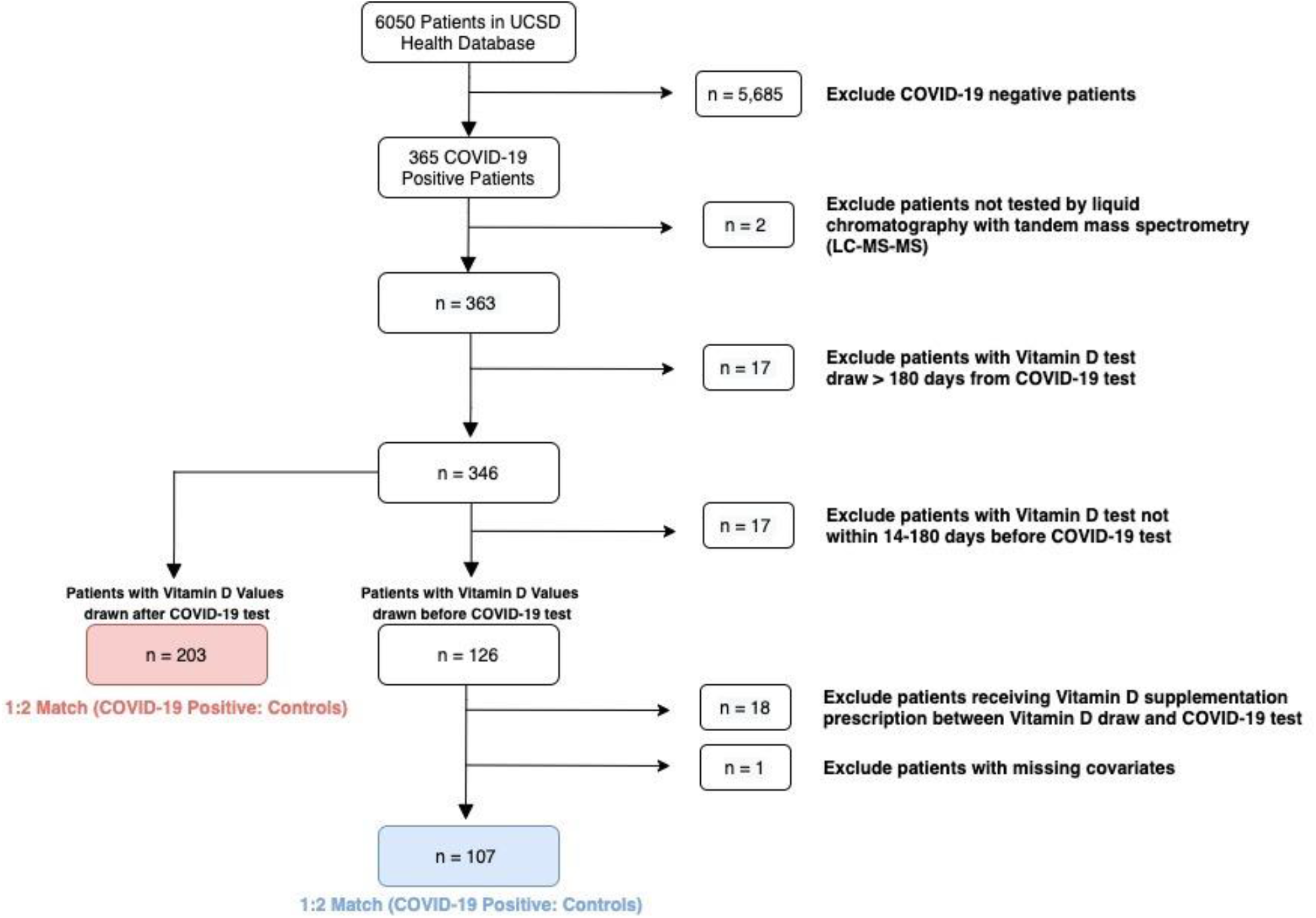
Flow diagram showing selection of study subjects and controls.

Following exclusions, there were 107 cases in the pre-diagnosis study and 203 cases in the post-diagnosis study. Cases in both studies were matched 1:2 to COVID-19 negative controls by age, sex, BMI, diagnosis of diabetes (ICD-10 codes E11.0-E11.9), diagnosis of hypertension (ICD-10 code I10), number of days between vitamin D draw and COVID-19 diagnosis, and meteorological season of 25(OH)D laboratory draw. Matching was performed using nearest-neighbor matching based on Mahalanobis distance. The distribution of serum 25(OH)D levels for each study group is shown in **Figure 2**.

**Figure 2.**
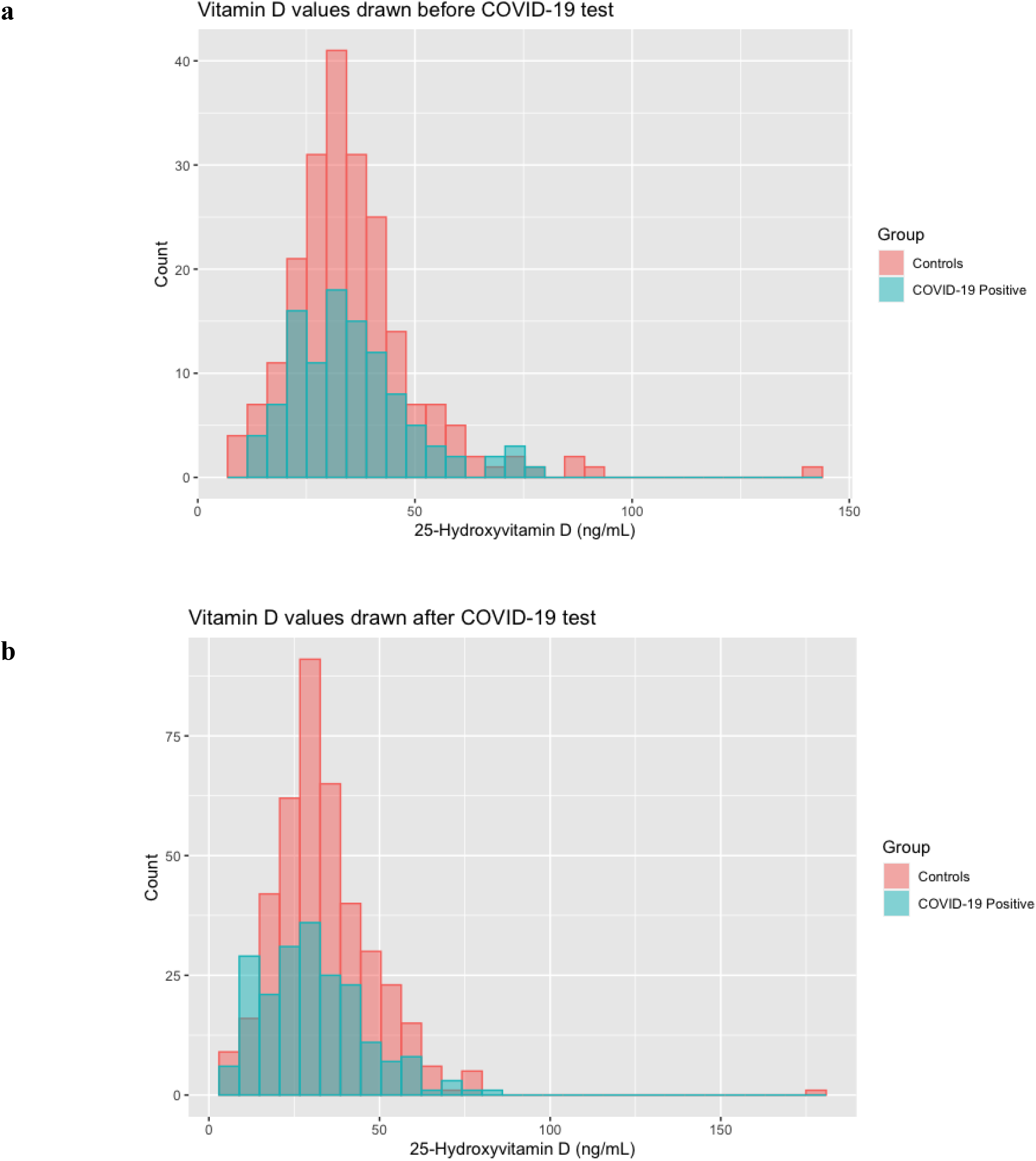
Distribution of serum 25(OH)D values for study populations. Histogram of (A) serum 25(OH)D values in subjects evaluated prior to COVID-19 diagnosis; (B) serum 25(OH)D values in subjects evaluated after COVID-19 diagnosis.

### Case-control and cohort studies

For the pre-diagnosis study, a case-control analysis was performed with 25(OH)D as the independent variable and COVID-19 infection status as the dependent variable. A conditional logistic regression model was performed to estimate the odds ratio (OR) and 95% confidence interval (95% CI) for serum 25(OH)D as a continuous variable. Results are reported as the change in odds of COVID-19 positivity for every 1 ng/mL increase in 25(OH)D.

For the post-diagnosis study, a historical cohort analysis was performed, in which COVID-19 infection status served as the binary independent variable to assess serum 25(OH)D levels as the outcome. An ordinary least squares (OLS) regression model was performed to estimate the change in serum 25(OH)D per ng/mL associated with COVID-19 positivity.

In a subset analysis of the second study, serum 25(OH)D levels in hospitalized COVID-19 patients were compared to COVID-19 negative hospitalized patients. Here, cases were defined as patients who had serum 25(OH)D drawn up to 180 days after inpatient admission due to COVID-19 (n=120). Matching criteria were identical to other analyses except for two changes: First, subjects were matched by the number of days between vitamin D draw and COVID-19 hospitalization, not COVID-19 diagnosis, and second, length of hospital stay was included as an additional matching covariate to promote comparison of hospitalized patients with similar disease severities^29^. An OLS regression model was performed to estimate the change in serum 25(OH)D per ng/mL associated with COVID-19 illness.

Analyses were performed using RStudio software, version 1.3.959. Continuous covariates are reported as mean ± standard deviation (SD) and compared using unpaired t-tests; categorical variables are reported as numbers and percentages and compared using chi-square tests. The significance level for all analyses was set to a two-sided p-value <0.05.

### Patient and Public Involvement

Subjects or the public were not involved in the design, or conduct, or reporting, or dissemination plans of this study.

## Data Availability

The datasets for this study are not publicly published due to presence of potential patient-identifiable information but will be made available from the corresponding author on reasonable request.

## ETHICS APPROVAL

This study was reviewed and approved by the UC San Diego Institutional Review Board (protocol #200768).

## DATA AVAILABILITY STATEMENT

The datasets for this study are not publicly published due to presence of potential patient-identifiable information but will be made available from the corresponding author on reasonable request.

